# Short- and longer-term all-cause mortality among SARS-CoV-2-infected persons and the pull-forward phenomenon in Qatar

**DOI:** 10.1101/2023.01.29.23285152

**Authors:** Hiam Chemaitelly, Jeremy Samuel Faust, Harlan M. Krumholz, Houssein H. Ayoub, Patrick Tang, Peter Coyle, Hadi M. Yassine, Asmaa A. Al Thani, Hebah A. Al-Khatib, Mohammad R. Hasan, Zaina Al-Kanaani, Einas Al-Kuwari, Andrew Jeremijenko, Anvar Hassan Kaleeckal, Ali Nizar Latif, Riyazuddin Mohammad Shaik, Hanan F. Abdul-Rahim, Gheyath K. Nasrallah, Mohamed Ghaith Al-Kuwari, Adeel A. Butt, Hamad Eid Al-Romaihi, Mohamed H. Al-Thani, Abdullatif Al-Khal, Roberto Bertollini, Laith J. Abu-Raddad

## Abstract

**Background:** Risk of short- and long-term all-cause mortality after a primary SARS-CoV-2 infection is inadequately understood.

**Methods:** A national, matched, retrospective cohort study was conducted in Qatar to assess the risk of all-cause mortality in the national cohort of people infected with SARS-CoV-2 compared with a reference national control cohort of uninfected persons. Associations were estimated using Cox proportional-hazards regression models.

**Results:** Among unvaccinated persons, within 90 days after primary infection, adjusted hazard ratio (aHR) comparing incidence of death in the primary-infection cohort with the infection-naïve cohort was 1.19 (95% CI: 1.02-1.39). The aHR was 1.34 (95% CI: 1.11-1.63) in persons more clinically vulnerable to severe COVID-19 and 0.94 (95% CI: 0.72-1.24) in those less clinically vulnerable to severe COVID-19. In subsequent follow-up, the aHR was 0.50 (95% CI: 0.37-0.68). The aHR was 0.41 (95% CI: 0.28-0.58) in months 3-7 after the primary infection and 0.76 (95% CI: 0.46-1.26) in subsequent months. The aHR was 0.37 (95% CI: 0.25-0.54) in persons more clinically vulnerable to severe COVID-19 and 0.77 (95% CI: 0.48-1.24) in those less clinically vulnerable to severe COVID-19. Among vaccinated persons, no evidence was found for differences in incidence of death in the primary-infection versus infection-naïve cohorts, even among persons more clinically vulnerable to severe COVID-19.

**Conclusions:** COVID-19 mortality in Qatar appears primarily driven by forward displacement of deaths of individuals with relatively short life expectancy and more clinically vulnerable to severe COVID-19. Vaccination negated the mortality displacement by preventing early deaths.

## Introduction

Severe acute respiratory syndrome coronavirus 2 (SARS-CoV-2) infection causes coronavirus disease 2019 (COVID-19) mortality.^1^ These deaths may occur as a result of factors related to the virus, the host, and available vaccinations and treatments. For example, COVID-19 death may occur because of the infection’s capacity to cause harm that results in death in otherwise healthy persons (viral virulence deaths). COVID-19 death may also occur because SARS-CoV-2 infection exacerbates the health condition of people who are vulnerable to short-term death independent of the virus because of severe underlying coexisting conditions or advanced age (forward displacement of mortality^2,3^). COVID-19 death may also happen because of long-term consequences of the acute SARS-CoV-2 infection, as systemic damage caused by the infection at time of infection may not be sufficient to cause immediate death, but could introduce health conditions either related to the acute infection or manifesting subsequently (Long COVID^4^) that can cause death at a later stage.

The extent to which each of these pathways contributes to COVID-19 mortality is unknown, whether in the total population or in specific population strata such as those defined by presence of vulnerability to severe COVID-19 or by vaccination status. Understanding the relative contribution of these pathways is critical to informing public health efforts on how best to deploy both pharmacologic and non-pharmacologic interventions that can favorably modify COVID-19 mortality. To address this knowledge gap, we investigated incidence of all-cause mortality, including COVID-19 mortality, after a primary SARS-CoV-2 infection in the Qatar national cohort of SARS-CoV-2 infected persons compared with a reference control cohort of infection-naïve persons from pandemic onset up to the present. We sought to elucidate the relative contribution of these pathways to the overall COVID-19 mortality.

## Methods

### Study population and data sources

This study was conducted on the population of Qatar using data between February 5, 2020, when the first SARS-CoV-2 infection was documented in this country, and November 14, 2022. We analyzed the national, federated, mortality database managed by the Hamad Medical Corporation, the national public healthcare provider in Qatar. The database includes all death records, including both deaths occurring at healthcare facilities and elsewhere (Section S1 of the Supplementary Appendix).

The study further analyzed the national, federated databases for COVID-19 laboratory testing, vaccination, and death, retrieved from the integrated, nationwide, digital-health information platform (Section S1). Databases include all SARS-CoV-2-related data with no missing information since the onset of the pandemic, including all polymerase chain reaction (PCR) tests and medically supervised rapid antigen tests (Section S2). SARS-CoV-2 testing in Qatar is done at large scale, and up to October 31, 2022, was mostly done for routine reasons such as for screening or travel-related purposes, with infections primarily diagnosed not because of appearance of symptoms, but because of routine testing.^5,6^ Qatar launched its COVID-19 vaccination program in December of 2020 using the BNT162b2 and mRNA-1273 vaccines.^7^ Detailed descriptions of Qatar’s population and of the national databases have been reported previously.^5,6,8-10^

### Study design and cohorts

A matched, retrospective, cohort study was conducted to compare incidence of all-cause mortality in the national cohort of persons who had a documented primary SARS-CoV-2 infection (designated the primary-infection cohort) to that in a national reference control cohort of infection-naïve persons (designated the infection-naïve cohort).

Primary infection was defined as the first record of a SARS-CoV-2-positive test regardless of symptoms. All-cause mortality was defined as any death, regardless of cause (including COVID-19), that occurred during follow-up.

### Cohorts matching, eligibility, and follow-up

Cohorts were matched exactly one-to-one by sex, 10-year age group, nationality, number of coexisting conditions (0, 1, 2, 3, 4, 5, or ≥6 coexisting conditions; Section S1), vaccination status (unvaccinated, 1, 2, 3, or 4 doses), and vaccine type (BNT162b2, mRNA-1273, or pediatric BNT162b2) to balance observed confounders between exposure groups that are potentially related to risks of mortality or infection.^8,11-14^ Matching was also done by calendar week of the test diagnosing the primary infection for persons in the primary-infection cohort and of a SARS-CoV-2-negative test for the infection-naïve cohort. That is, persons who had the primary infection diagnosed in a specific calendar week were matched to unique infection-naïve persons who had a record of a SARS-CoV-2-negative test in that same calendar week. This was done to ensure that those in the infection-naïve cohort were not infected at the time of recruitment, to control for time-variable differences in mortality risk, and to ensure that matched pairs were present in Qatar in the same period.

Cohorts were also matched exactly by SARS-CoV-2 testing method (PCR versus rapid antigen testing) and by reason for testing (Section S1) to control for potential differences in presence of testing modalities between cohorts.

Persons were eligible for inclusion in the primary-infection cohort if they had a record for a SARS-CoV-2-positive test, but no record for vaccine doses mixing different vaccines by the start of follow-up. Persons were eligible for inclusion in the infection-naïve cohort if they had a record for at least one SARS-CoV-2-negative test. Persons with a record of vaccination with a non-mRNA vaccine or an unspecified vaccine type were excluded.

Matching was performed iteratively such that persons in the infection-naïve control cohort were alive and had no record, by the start of follow-up, for a SARS-CoV-2-positive test, or for vaccine doses mixing different vaccines, or for a new vaccine dose between the time of the SARS-CoV-2-negative test and the start date of follow-up.

Follow-up was from 1 day before the primary infection to account for situations where testing was done immediately post-mortem (to determine cause of death), through the end of the study (November 14, 2022). Some persons contributed follow-up time first in the infection-naïve cohort, while matched to persons with primary infections, and subsequently contributed data to the primary-infection cohort after infection (at which point they were matched to persons who were still infection-naïve). Matching was iterated with as many replications as needed until exhaustion (i.e., no more matched pairs could be identified).

For exchangeability,^10,15^ both members of each matched pair were censored as soon as the person in the primary-infection cohort was reinfected, or the person in the infection-naïve cohort was infected, or one of them received a new vaccine dose (change in vaccination status). Reinfection was defined as a documented infection ≥90 days after an earlier infection, to avoid misclassification of prolonged SARS-CoV-2-positivity as reinfection.^16,17^ Accordingly, individuals were followed up until the first of any of the following events: a documented reinfection (with matched pair censoring), a documented infection (with matched pair censoring), a change in vaccination status (with matched-pair censoring), death from any cause, or administrative end of follow-up (November 14, 2022).

### Classification of COVID-19 death

Classification of COVID-19 death followed World Health Organization (WHO) guidelines^18^ (Section S1). Assessments were made by trained medical personnel independent of study investigators and using individual chart reviews, as part of a national protocol applied to every deceased patient since pandemic onset.

### Oversight

The institutional review boards at Hamad Medical Corporation and Weill Cornell Medicine– Qatar approved this retrospective study with a waiver of informed consent. The study was reported according to the Strengthening the Reporting of Observational Studies in Epidemiology (STROBE) guidelines (Table S1). The authors vouch for the accuracy and completeness of the data and for the fidelity of the study to the protocol. Data used in this study are the property of the Ministry of Public Health of Qatar and were provided to the researchers through a restricted-access agreement for preservation of confidentiality of patient data. The funders had no role in the study design; the collection, analysis, or interpretation of the data; or the writing of the manuscript.

### Statistical analysis

Eligible and matched cohorts were described using frequency distributions and measures of central tendency and were compared using standardized mean differences (SMDs). An SMD of ≤0.1 indicated adequate matching.^19^ For each of unvaccinated and vaccinated persons, two analyses were conducted to compare incidence of all-cause mortality in the primary-infection and infection-naïve cohorts during the first 90 days after the primary infection (Acute SARS-CoV-2 Infection Mortality Analysis) and from 91 days and thereafter (Post-acute SARS-CoV-2 Mortality Analysis).

Cumulative incidence of death (defined as proportion of persons at risk, whose primary endpoint during follow-up was death) was estimated using the Kaplan-Meier estimator method. Incidence rate of death in each of unvaccinated and vaccinated persons of each cohort, defined as number of deaths divided by number of person-years contributed by all individuals in the cohort, was estimated, with the corresponding 95% confidence interval (CI), using a Poisson log-likelihood regression model with the Stata 17.0 *stptime* command.

Overall hazard ratios, comparing incidence of death in the cohorts and corresponding 95% CIs, were calculated using Cox regression adjusted for the matching factors with the Stata 17.0 *stcox* command. The overall hazard ratios provided a weighted average of the time-varying hazard ratios.^20^ Adjusted hazard ratios were also estimated for different time intervals after start of follow-up using separate Cox regressions with “failures” restricted to specific time intervals.

Schoenfeld residuals and log-log plots for survival curves were used to examine the proportional-hazards assumption. CIs were not adjusted for multiplicity; thus, they should not be used to infer definitive differences between groups. Interactions were not considered except in sensitivity analysis.

Adjusted hazard ratios were estimated for subgroups of the study cohorts including persons who are less or more clinically vulnerable to severe COVID-19. Persons less clinically vulnerable to severe COVID-19 were defined as those <50 years of age and with one or no coexisting

conditions.^21^ Persons more clinically vulnerable to severe COVID-19 were defined as those ≥50 years of age, or <50 years of age but with ≥2 coexisting conditions.^21^ The conceptual approach in this study was to construct the matched cohorts in a manner that allows disaggregation of the full cohorts into separate sub-studies for each of unvaccinated and vaccinated persons and assess study outcomes for these subgroups. A disadvantage is that this may entail different population compositions other than the ones of interest. In a sensitivity analysis to investigate whether this may have affected study results, study outcomes for these subgroups were calculated instead using interaction terms with the Cox interaction model applied to the full cohorts.

Another sensitivity analysis was conducted by restricting the study to only Qataris, that is excluding all expatriates, to assess whether travel or leaving the country for expatriates may have affected study results. Statistical analyses were performed using Stata/SE version 17.0 (Stata Corporation, College Station, TX, USA).

## Results

Figure S1 shows the study population selection process. Table 1 describes baseline characteristics of the full and matched cohorts. Matched cohorts each included 685,871 persons. The study population is representative of the population of Qatar (Table S2).

**Table 1.** Baseline characteristics of full and matched cohorts.

| Characteristics | Full eligible cohorts |  |  | Matched cohorts* |  |  |
| --- | --- | --- | --- | --- | --- | --- |
|  | Primary-infection | Infection-naïve | SMD† | Primary-infection | Infection-naïve | SMD† |
|  | N=890,705 | N=3,965,765 |  | N=685,871 | N=685,871 |  |
| Median age (IQR)—years | 33 (24-41) | 32 (25-40) | 0.01‡ | 32 (23-39) | 32 (23-39) | 0.00‡ |
| Age—years |  |  |  |  |  |  |
| 0-9 years | 86,986 (9.8) | 359,438 (9.1) |  | 73,165 (10.7) | 73,165 (10.7) |  |
| 10-19 years | 84,120 (9.4) | 272,855 (6.9) |  | 63,764 (9.3) | 63,764 (9.3) |  |
| 20-29 years | 183,616 (20.6) | 973,741 (24.6) |  | 151,561 (22.1) | 151,561 (22.1) |  |
| 30-39 years | 282,025 (31.7) | 1,280,464 (32.3) | 0.13 | 226,172 (33.0) | 226,172 (33.0) | 0.00 |
| 40-49 years | 158,757 (17.8) | 678,087 (17.1) |  | 116,971 (17.1) | 116,971 (17.1) |  |
| 50-59 years | 66,108 (7.4) | 277,233 (7.0) |  | 41,183 (6.0) | 41,183 (6.0) |  |
| 60-69 years | 21,514 (2.4) | 94,683 (2.4) |  | 10,418 (1.5) | 10,418 (1.5) |  |
| 70+ years | 7,579 (0.9) | 29,264 (0.7) |  | 2,637 (0.4) | 2,637 (0.4) |  |
| Sex |  |  |  |  |  |  |
| Male | 558,518 (62.7) | 2,805,558 (70.7) | 0.17 | 445,511 (65.0) | 445,511 (65.0) | 0.00 |
| Female | 332,187 (37.3) | 1,160,207 (29.3) |  | 240,360 (35.0) | 240,360 (35.0) |  |
| Nationality§ |  |  |  |  |  |  |
| Bangladeshi | 50,496 (5.7) | 289,261 (7.3) |  | 41,106 (6.0) | 41,106 (6.0) |  |
| Egyptian | 47,114 (5.3) | 190,814 (4.8) |  | 34,771 (5.1) | 34,771 (5.1) |  |
| Filipino | 88,481 (9.9) | 294,798 (7.4) |  | 70,056 (10.2) | 70,056 (10.2) |  |
| Indian | 200,810 (22.6) | 1,111,610 (28.0) |  | 175,997 (25.7) | 175,997 (25.7) |  |
| Nepalese | 61,707 (6.9) | 374,369 (9.4) | 0.37 | 50,905 (7.4) | 50,905 (7.4) | 0.00 |
| Pakistani | 37,770 (4.2) | 231,607 (5.8) |  | 29,990 (4.4) | 29,990 (4.4) |  |
| Qatari | 170,274 (19.1) | 319,522 (8.1) |  | 134,475 (19.6) | 134,475 (19.6) |  |
| Sri Lankan | 22,739 (2.6) | 131,015 (3.3) |  | 17,499 (2.6) | 17,499 (2.6) |  |
| Sudanese | 23,152 (2.6) | 81,538 (2.1) |  | 15,957 (2.3) | 15,957 (2.3) |  |
| Other nationalities¶ | 188,162 (21.1) | 941,231 (23.7) |  | 115,115 (16.8) | 115,115 (16.8) |  |
| Coexisting conditions |  |  |  |  |  |  |
| None | 688,703 (77.3) | 3,483,150 (87.8) |  | 562,885 (82.1) | 562,885 (82.1) |  |
| 1 | 111,556 (12.5) | 274,952 (6.9) |  | 76,009 (11.1) | 76,009 (11.1) |  |
| 2 | 46,373 (5.2) | 108,253 (2.7) |  | 27,069 (4.0) | 27,069 (4.0) |  |
| 3 | 19,641 (2.2) | 45,366 (1.1) | 0.28 | 9,545 (1.4) | 9,545 (1.4) | 0.00 |
| 4 | 10,925 (1.2) | 25,145 (0.6) |  | 4,735 (0.7) | 4,735 (0.7) |  |
| 5 | 6,538 (0.7) | 14,363 (0.4) |  | 2,542 (0.4) | 2,542 (0.4) |  |
| 6+ | 6,969 (0.8) | 14,536 (0.4) |  | 3,086 (0.5) | 3,086 (0.5) |  |
| Vaccination status** |  |  |  |  |  |  |
| Unvaccinated | 545,756 (61.3) | 2,665,308 (67.2) |  | 460,620 (67.2) | 460,620 (67.2) |  |
| 1 dose | 185,93 (2.1) | 53,244 (1.3) |  | 10,366 (1.5) | 10,366 (1.5) |  |
| 2 doses | 257,358 (28.9) | 1,006,540 (25.4) | 0.13 | 168,939 (24.6) | 168,939 (24.6) | 0.00 |
| 3 doses | 68,602 (7.7) | 238,708 (6.0) |  | 45,900 (6.7) | 45,900 (6.7) |  |
| 4 doses | 396 (0.04) | 1,965 (0.05) |  | 46 (0.01) | 46 (0.01) |  |
| Vaccine type |  |  |  |  |  |  |
| Unvaccinated | 545,756 (61.3) | 2,665,308 (67.2) |  | 460,620 (67.2) | 460,620 (67.2) |  |
| BNT162b2 | 251,345 (28.2) | 802,053 (20.2) | 0.19 | 165,657 (24.2) | 165,657 (24.2) | 0.00 |
| mRNA-1273 | 92,114 (10.3) | 487,864 (12.3) |  | 58,624 (8.6) | 58,624 (8.6) |  |
| Pediatric BNT162b2 | 1,490 (0.2) | 10,540 (0.3) |  | 970 (0.1) | 970 (0.1) |  |
| Testing method |  |  |  |  |  |  |
| PCR | 700,342 (78.6) | 2,889,278 (72.9) | 0.13 | 551,228 (80.4) | 551,228 (80.4) | 0.00 |
| Rapid antigen | 190,363 (21.4) | 1,076,487 (27.1) |  | 134,643 (19.6) | 134,643 (19.6) |  |
| Reason for testing |  |  |  |  |  |  |
| Clinical suspicion | 253,259 (28.4) | 290,431 (7.3) |  | 164,763 (24.0) | 164,787 (24.0) |  |
| Contact tracing | 131,087 (14.7) | 203,717 (5.1) |  | 104,321 (15.2) | 104,331 (15.2) |  |
| Survey | 89,513 (10.1) | 444,064 (11.2) |  | 77,090 (11.2) | 77,146 (11.3) |  |
| Individual request | 53,690 (6.0) | 282,014 (7.1) |  | 40,193 (5.9) | 40,168 (5.9) |  |
| Pre-travel | 92,031 (10.3) | 739,057 (18.6) | 0.84 | 76,575 (11.2) | 76,613 (11.2) | 0.00 |
| Port of entry | 96,420 (10.8) | 1,252,304 (31.6) |  | 91,876 (13.4) | 91,872 (13.4) |  |
| Healthcare routine testing | 31,715 (3.6) | 138,225 (3.5) |  | 24,505 (3.6) | 24,534 (3.6) |  |
| Other | 9,169 (1.0) | 36,721 (0.9) |  | 6,756 (1.0) | 6,758 (1.0) |  |
| Unspecified | 133,821 (15.0) | 579,232 (14.6) |  | 99,792 (14.6) | 99,542 (14.5) |  |

IQR denotes interquartile range, SARS-CoV-2 severe acute respiratory syndrome coronavirus 2, and SMD standardized mean difference.
†Cohorts were matched exactly one-to-one by sex, 10-year age group, nationality, number of coexisting conditions, vaccination status, vaccine type, SARS-CoV-2 testing method, reason for testing, and calendar week of testing.
‡SMD is the difference in the mean of a covariate between groups divided by the pooled standard deviation. An SMD ≤0.1 indicates adequate matching.
§SMD is for the mean difference between groups divided by the pooled standard deviation.
¶Nationalities were chosen to represent the most populous groups in Qatar.
\*\*These comprise up to 183 other nationalities in the unmatched cohort and 144 other nationalities in the matched cohort.
\*\*Ascertained at the time of the SARS-CoV-2 test.

### Unvaccinated population

#### Acute SARS-CoV-2 Infection Mortality Analysis

In this analysis, the median follow-up was 91 days (interquartile range (IQR), 65-91 days) (Figure 1A). During follow-up, 342 deaths were recorded in the primary-infection cohort compared to 288 deaths in the infection-naïve cohort (Figure S1 and Table 2). Of the 342 deaths in the primary-infection cohort, 223 were COVID-19 deaths. Cumulative incidence of death was 0.085% (95% CI: 0.076-0.095%) in the primary-infection cohort and 0.072% (95% CI: 0.064-0.081%) in the infection-naïve cohort, 91 days after the start of follow-up (Figure 1A).

**Table 2.** Adjusted hazard ratio for all-cause death in the matched primary-infection and infection-naïve cohorts for each of unvaccinated and vaccinated persons.

|  | Unvaccinated |  | Vaccinated |  |
| --- | --- | --- | --- | --- |
| Epidemiological measures | Primary-infection cohort* | Control cohort* | Primary-infection cohort* | Control cohort* |
| Acute SARS-CoV-2 Infection Mortality Analysis |  |  |  |  |
| Sample size | 460,620 | 460,620 | 225,251 | 225,251 |
| Total follow-up time (person-years) | 94,267 | 94,271 | 40,426 | 40,424 |
| Number of deaths during follow-up | 342 | 288 | 59 | 74 |
| Incidence rate of death (per 1,000 person-years; 95% CI) | 3.63 (3.26 to 4.03) | 3.06 (2.72 to 3.43) | 1.46 (1.13 to 1.88) | 1.83 (1.46 to 2.30) |
| Unadjusted hazard ratio for death (95% CI) | 1.17 (1.00 to 1.36) |  | 0.80 (0.57 to 1.12) |  |
| Adjusted hazard ratio for death (95% CI)<sup>†</sup> | 1.19 (1.02 to 1.39) |  | 0.74 (0.50 to 1.09) |  |
| Post-acute SARS-CoV-2 Infection Mortality Analysis |  |  |  |  |
| Sample size | 315,489 | 315,506 | 131,960 | 131,942 |
| Total follow-up time (person-years) | 187,359 | 187,319 | 56,521 | 56,509 |
| Number of deaths during follow-up | 72 | 142 | 45 | 44 |
| Incidence rate of death (per 1,000 person-years; 95% CI) | 0.38 (0.31 to 0.48) | 0.76 (0.64 to 0.89) | 0.80 (0.59 to 1.07) | 0.78 (0.58 to 1.05) |
| Unadjusted hazard ratio for death (95% CI) | 0.51 (0.38 to 0.68) |  | 1.02 (0.67 to 1.55) |  |
| Adjusted hazard ratio for death (95% CI)<sup>†</sup> | 0.50 (0.37 to 0.66) |  | 1.10 (0.68 to 1.78) |  |

CI denotes confidence interval.
\*Each person in the primary-infection cohort was matched exactly one-to-one by sex, 10-year age group, nationality, number of coexisting conditions, vaccination status, vaccine type, SARS-CoV-2 testing method, reason for testing, and calendar week of testing to a person with a SARS-CoV-2-negative test who, by the start of the follow-up, was alive, infection free, and did not receive vaccine doses with different vaccines, or a new vaccine dose between the SARS-CoV-2-negative test and the start date of follow-up.
<sup>†</sup>Cox regression analysis adjusted for sex, 10-year age group, 10 nationality groups, number of coexisting conditions, vaccination status, vaccine type, SARS-CoV-2 testing method, reason for testing, and calendar week of testing.

**Figure 1.**
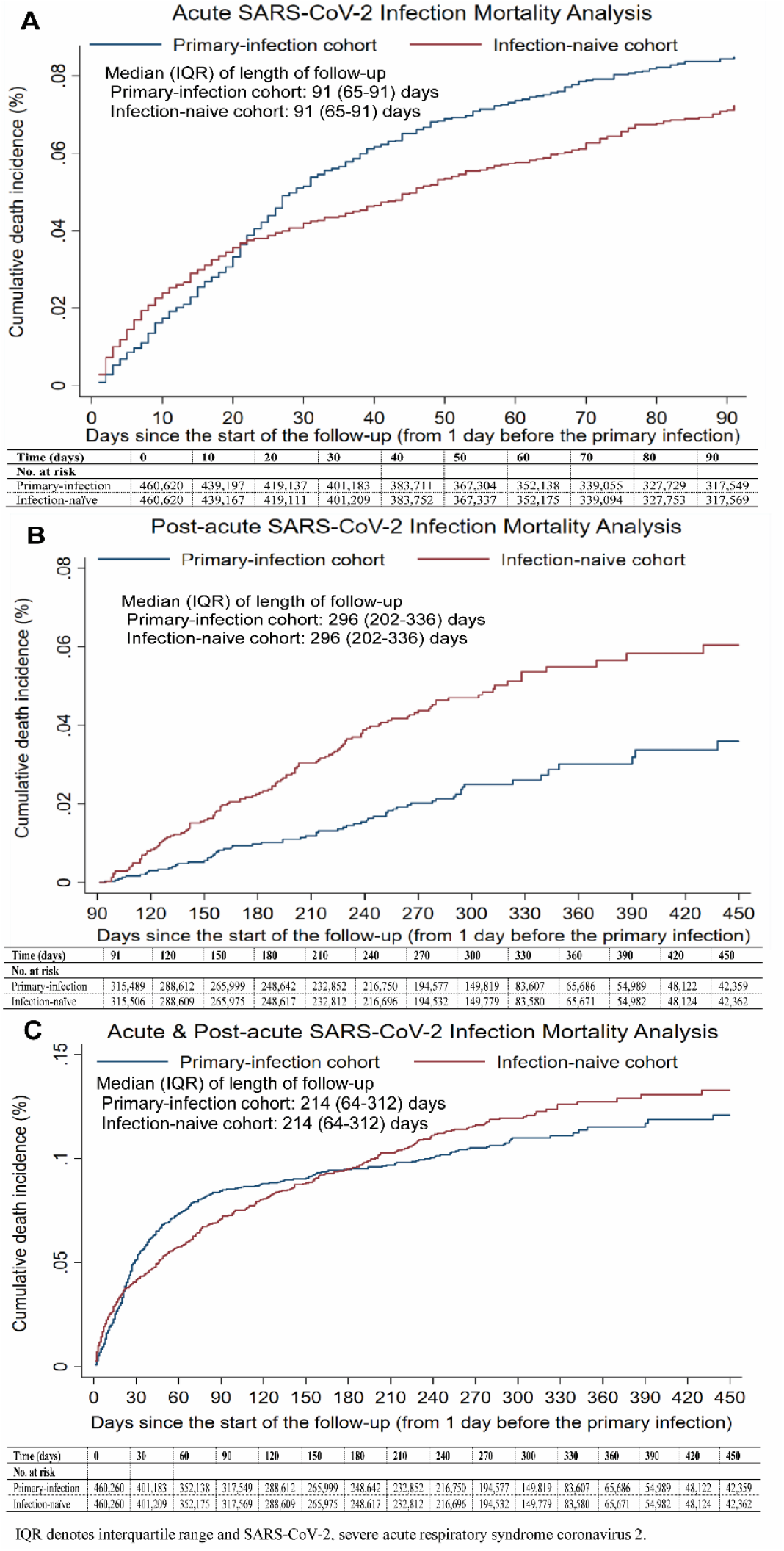
Cumulative incidence of all-cause death among unvaccinated persons of the matched primary-infection and infection-naïve cohorts in the A) Acute SARS-CoV-2 Infection Mortality Analysis, B) Post-acute SARS-CoV-2 Infection Mortality Analysis, and C) Combined Acute & Post-acute SARS-CoV-2 Infection Mortality Analysis

Adjusted hazard ratio (aHR) comparing incidence of death in the unvaccinated primary-infection cohort to the unvaccinated infection-naïve cohort during the acute phase was 1.19 (95% CI: 1.02-1.39; Table 2 and Figure 2A). Subgroup analyses estimated the aHR at 1.34 (95% CI: 1.11-1.63) in persons more clinically vulnerable to severe COVID-19 and 0.94 (95% CI: 0.72-1.24) in those less clinically vulnerable to severe COVID-19 (Figure 3A and Table S3A).

**Figure 2.**
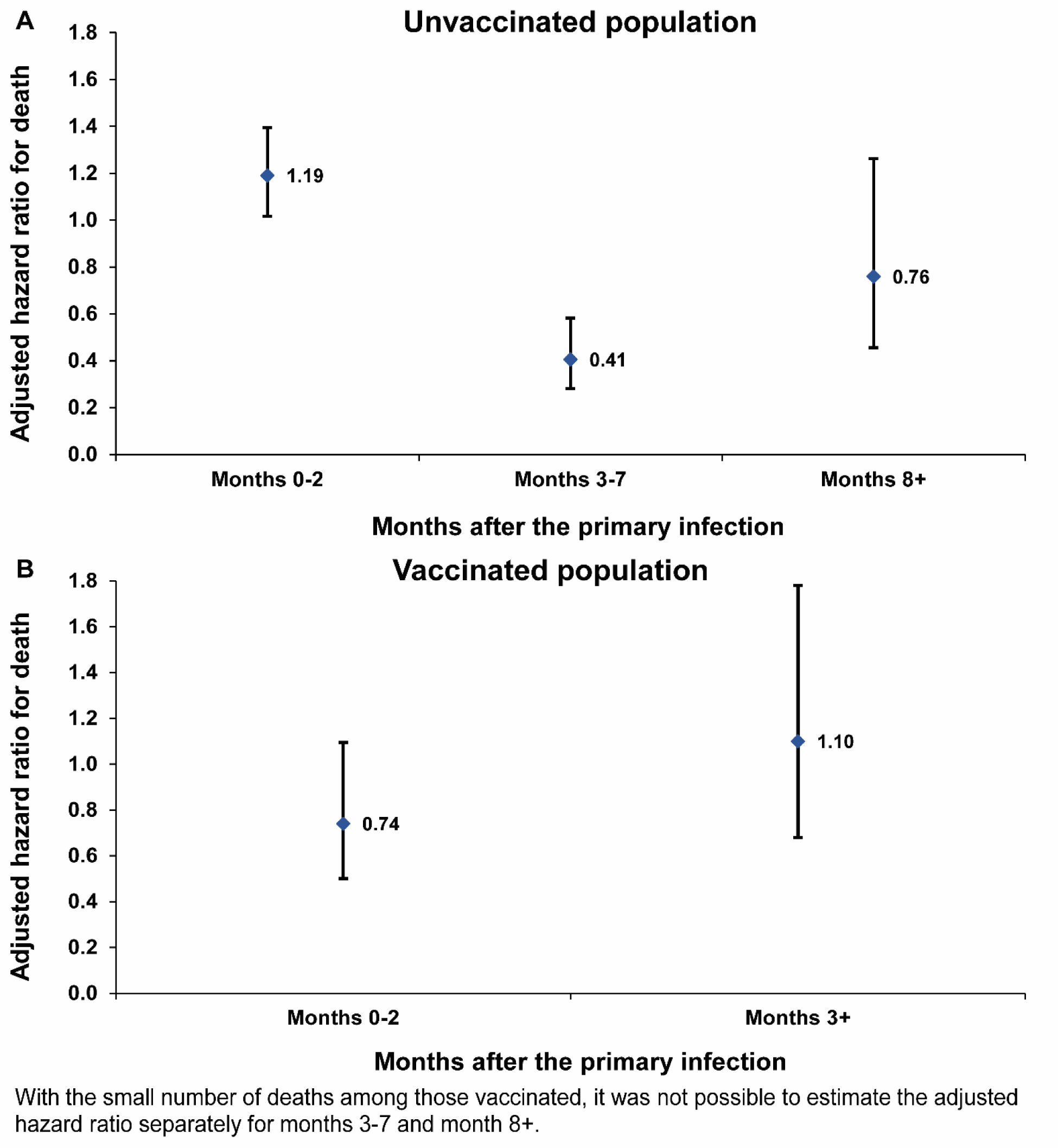
Adjusted hazard ratios for all-cause death by time since the start of follow-up in the matched primary-infection and infection-naïve cohorts for each of A) unvaccinated and vaccinated persons.

**Figure 3.**
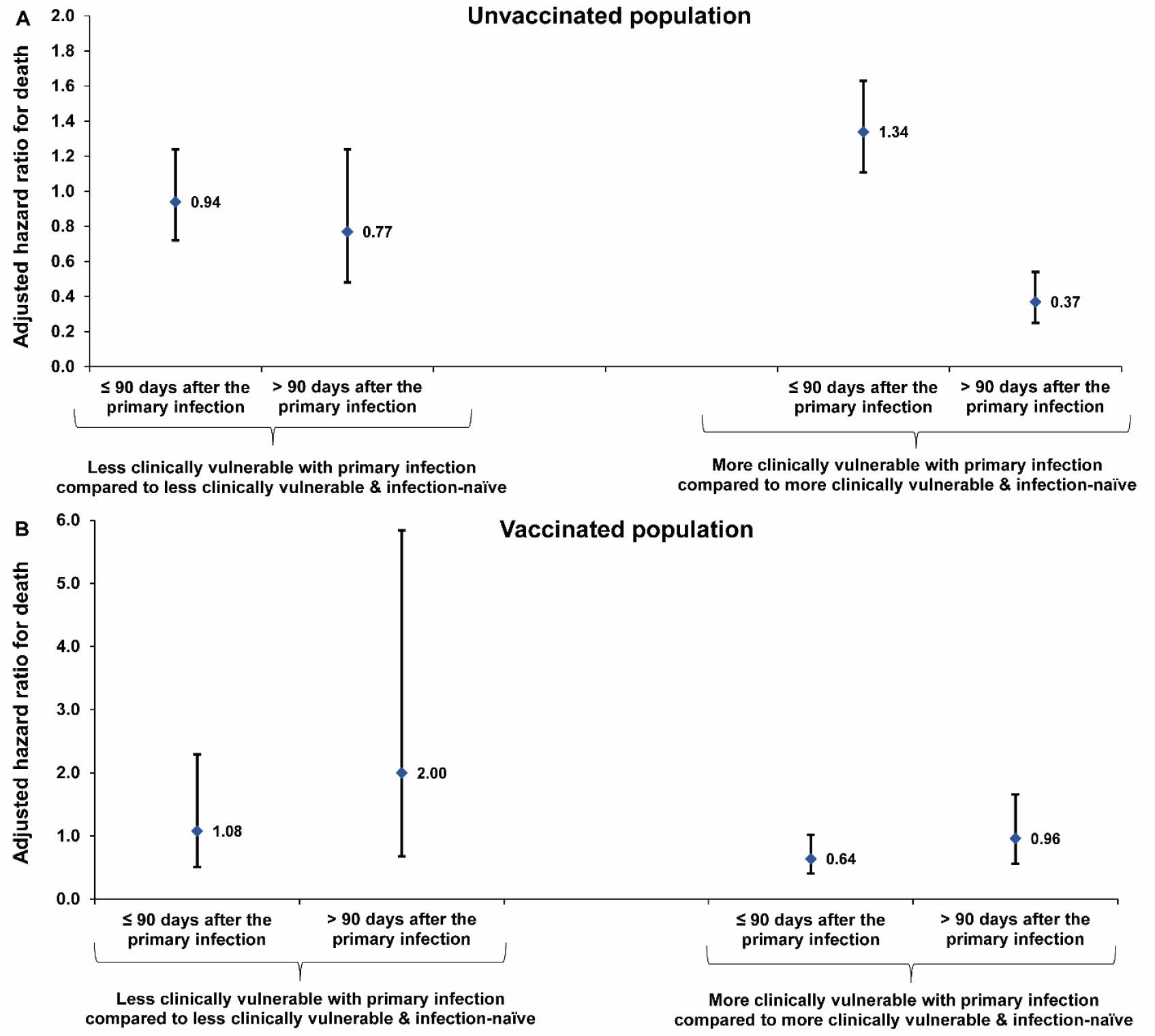
Adjusted hazard ratio for all-cause death in the A) unvaccinated and B) vaccinated populations by clinical vulnerability to severe COVID-19 in the Acute SARS-CoV-2 Infection Mortality Analysis (≤90 days after the primary infection) and in the Post-acute SARS-CoV-2 Infection Mortality Analysis (>90 days after the primary infection).

#### Post-acute SARS-CoV-2 Infection Mortality Analysis

In this analysis, the median follow-up was 296 days (IQR, 202-336 days) for each of the cohorts (Figure 1B). During follow-up, 72 deaths were recorded in the primary-infection cohort compared to 142 in the infection-naïve cohort (Figure S1 and Table 2). Of the 72 deaths in the primary-infection cohort, 5 were COVID-19 deaths related to the original primary infection.

Cumulative incidence of death was 0.036% (95% CI: 0.027-0.048%) in the primary-infection cohort and 0.060% (95% CI: 0.050-0.074%) in the infection-naïve cohort, 450 days after the start of follow-up (Figure 1B).

The aHR comparing incidence of death in the unvaccinated primary-infection cohort to the unvaccinated infection-naïve cohort during the post-acute phase was 0.50 (95% CI: 0.37-0.68; Table 2). The aHR was 0.41 (95% CI: 0.28-0.58) in months 3-7 after the primary infection and increased to 0.76 (95% CI: 0.46-1.26) in subsequent months (Figure 2A). The aHR during the post-acute phase was 0.37 (95% CI: 0.25-0.54) in persons more clinically vulnerable to severe COVID-19 and 0.77 (95% CI: 0.48-1.24) in those less clinically vulnerable to severe COVID-19 (Figure 3A and Table S3A).

#### Combined Acute and Post-acute SARS-CoV-2 Infection Mortality Analysis

Figure 1C shows cumulative incidence of death in the unvaccinated population covering the entire time of follow-up, including both acute and post-acute phases. The aHR comparing incidence of death in the unvaccinated primary-infection cohort to the unvaccinated infection-naïve cohort was 0.97 (95% CI: 0.84-1.11). The aHR was 1.00 (95% CI: 0.85-1.19) in persons more clinically vulnerable to severe COVID-19 and 0.90 (95% CI: 0.71-1.14) in persons less clinically vulnerable to severe COVID-19 (Table S3A).

### Vaccinated population

In the Acute SARS-CoV-2 Infection Mortality Analysis (Figure 4A), the aHR comparing incidence of death in the vaccinated primary-infection cohort to the vaccinated infection-naïve cohort was 0.74 (95% CI: 0.50-1.09; Table 2 and Figure 2B). Subgroup analyses estimated the aHR at 0.64 (95% CI: 0.41-1.02) in persons more clinically vulnerable to severe COVID-19 and 1.08 (95% CI: 0.51-2.29) in those less clinically vulnerable to severe COVID-19 (Figure 3B and Table S3B).

**Figure 4.**
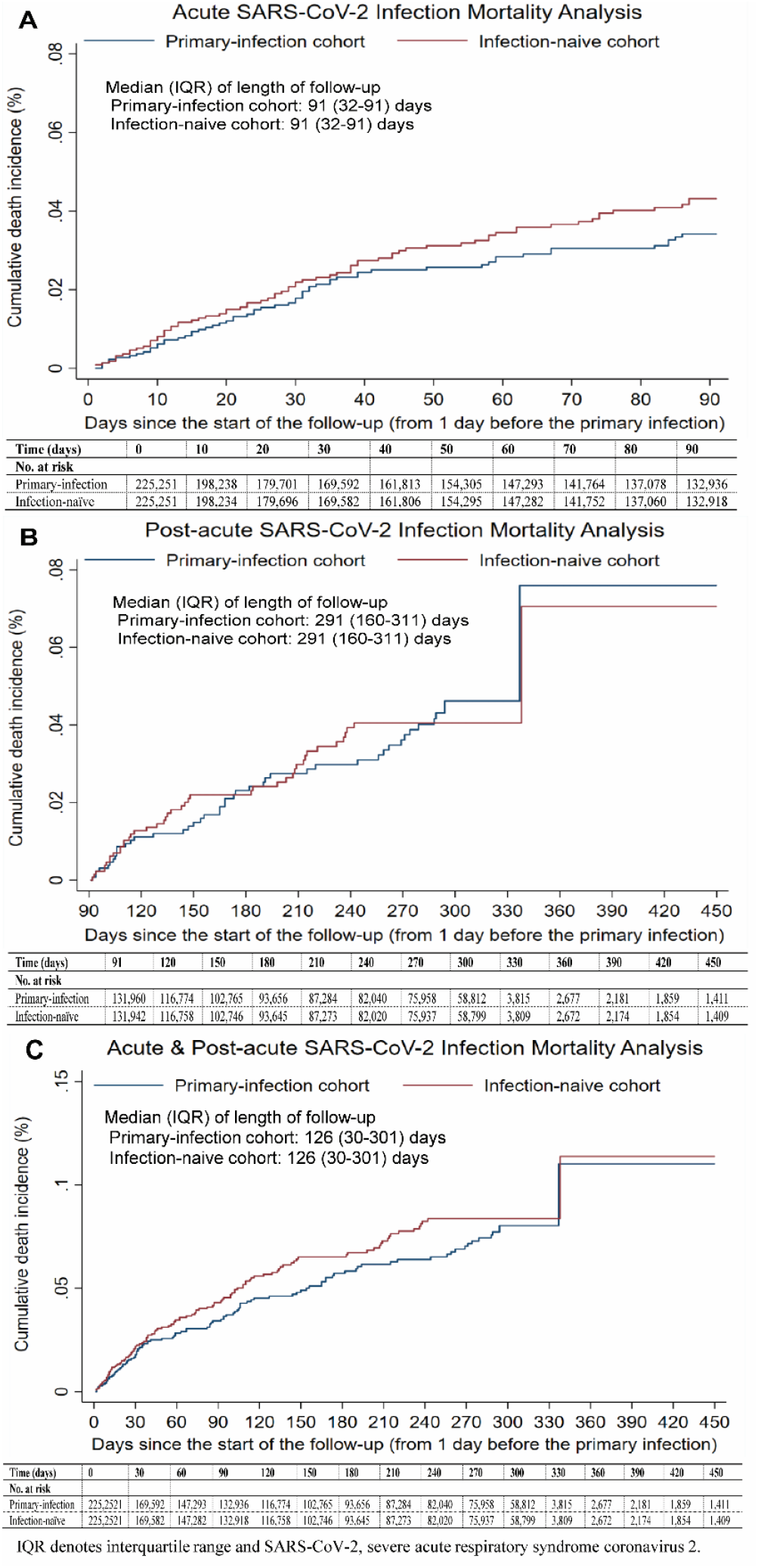
Cumulative incidence of all-cause death among vaccinated persons of the matched primary-infection and infection-naïve cohorts in the A) Acute SARS-CoV-2 Infection Mortality Analysis, B) Post-acute SARS-CoV-2 Infection Mortality Analysis, and Combined Acute & Post-acute Infection Mortality Analysis.

In the Post-acute SARS-CoV-2 Infection Mortality Analysis (Figure 4B), the aHR comparing incidence of death in the vaccinated primary-infection cohort to the vaccinated infection-naïve cohort was 1.10 (95% CI: 0.68-1.78; Table 2 and Figure 2B). Subgroup analyses estimated the aHR at 0.96 (95% CI: 0.56-1.66) in persons more clinically vulnerable to severe COVID-19 and 2.00 (95% CI: 0.68-5.84) in those less clinically vulnerable to severe COVID-19 (Figure 3B and Table S3B).

Figure 4C shows cumulative incidence of death in the vaccinated population in the Combined Acute and Post-acute SARS-CoV-2 Infection Mortality Analysis, covering the entire period of follow-up. aHR was 0.87 (95% CI: 0.64-1.18). The aHR was 0.76 (95% CI: 0.53-1.07) and 1.33 (95% CI: 0.72-2.46) in persons more and less clinically vulnerable to severe COVID-19, respectively (Table S3B).

### Additional and sensitivity analyses

In the analysis combining acute and post-acute phases and unvaccinated and vaccinated persons, the aHR comparing incidence of death in the primary-infection cohort to the infection-naïve cohort was 0.95 (95% CI: 0.84-1.07; Table S4).

The sensitivity analysis calculating study outcomes through interaction terms with the interaction model applied to the full cohorts produced similar results to main analysis (Table S5A). The sensitivity analysis calculating study outcomes only for Qataris also generated similar results to main analysis (Table S5B).

## Discussion

The findings are consistent with COVID-19 mortality in Qatar being driven primarily by forward displacement of deaths of individuals with relatively short life expectancy. Incidence of death among those infected versus those infection-naïve demonstrated a V-shape (Figure 2), characteristic of forward mortality displacement.^2,3^ Incidence of all-cause mortality was higher among those infected in the short-term (excess mortality), lower among those infected in the intermediate term (deficit mortality), with no evidence for differences in incidence of death observed by the end of the study period. This is supported by a recent observation of the temporary occurrence of deficit mortality after large COVID-19 waves.^22^ The pull forward effect was observed in the overall cohort, but markedly pronounced among those more clinically vulnerable to severe COVID-19. Meanwhile, no such effect was observed among those less clinically vulnerable. In that subgroup, the hazard ratio for death comparing the infected to uninfected was approximately 1, during both the acute and post-acute phases of follow-up, further supporting mortality displacement among the clinically vulnerable as the main driver of COVID-19 mortality.

These findings do not diminish the importance of COVID-19 deaths in otherwise healthy persons or due to Long COVID. Such deaths have been documented globally. However, there was no discernable effect for these deaths in Qatar’s population, perhaps because of insufficient statistical power to measure small effects compared to the large effect of forward displacement of deaths, even in such large cohort study.

Vaccination negated the infection’s pull forward (mortality displacement) effect by preventing early deaths. The hazard ratio for death was approximately 1 among vaccinated persons during both the acute and post-acute phases of follow-up, regardless of the clinical vulnerability to severe COVID-19. This revealed the occurrence of a theoretically expected (though potentially counterintuitive) finding; the incidence of all-cause death would be expected to be higher among vaccinated compared to unvaccinated persons at this later stage of the pandemic. This is because clinically vulnerable vaccinated persons had higher survival probabilities through repeated waves of infection and thus, unlike unvaccinated persons with similarly elevated baseline mortality risks, were able to survive long enough to contribute follow-up time to the analysis.

These findings may explain the low rate of COVID-19 mortality in Qatar, one of the lowest worldwide.^23^ As of January 29, 2023, 686 COVID-19 deaths have been recorded in this country of three million people;^24^ <0.1% of documented infections ended in COVID-19 death. About 60% of Qatar’s population are craft and manual workers (CMW)^13,14^ who are typically single men, 20-49 years of age.^25^ CMW are healthy by recruitment (healthy worker effect^26,27^) and have lower levels of comorbidities such as diabetes and obesity;^28^ that is a population not likely to be affected by the pull forward effect.

This study has limitations. Documented COVID-19 deaths may not include all deaths that occurred because of COVID-19.^3,29^ Some COVID-19 deaths may not have been confirmed due to insufficient information to confirm COVID-19 as the cause of death.^3,29^ The differences in incidence rate of non-COVID-19 death between the primary-infection and infection-naïve cohorts in the acute phase of follow-up, which coincided with calendar times of high infection incidence during waves, suggest existence of COVID-19-associated deaths among those in the “infection-naïve” cohort that were never detected or confirmed.

All-cause mortality appeared qualitatively elevated among uninfected persons during the first 0-3 weeks after their matched pairs were infected. This observation may have risen because of differences in the risk of non-COVID-19 death between those recently diagnosed with the infection and uninfected persons. Isolation of infected persons in Qatar was enforced through mandatory requirements and isolation facilities. The reduced mobility of infected persons following diagnosis of infection should have reduced their risk of incidental causes of death, such as motor-vehicle road injuries, a leading cause of death that contributes ∼10% of all deaths in Qatar.^30-32^ An additional explanation is the selection of controls among those with a negative test. A recent negative test is perhaps a proxy of recent activity and mobility, and thus of higher likelihood of incidental death. Persons who are less active and staying at home are less likely to do a routine or screening test or to experience an incidental death.

Primary infections were determined based on records of documented infections, but other infections may have occurred and gone undocumented, for example due to tests being administered prematurely (i.e., during the viral incubation period). This may also explain the qualitatively elevated risk of death among uninfected persons during the first 0-3 weeks of follow-up. Tests performed on persons with known exposure may have in some instances been administered too early, before viral loads were sufficient to mount positive tests.

Improvements in case management, use of antivirals, and introduction of omicron with its lower infection severity^33,34^ reduced COVID-19 mortality over time. This may have affected the observed trends; most of COVID-19 mortality effects were driven by incidence during the pre-omicron waves. The study analyzed all deaths that occurred in Qatar, but some deaths may have occurred outside Qatar, while expatriates were traveling abroad or if they left the country because of end of employment after initiation of follow-up. Travel data were not available to be incorporated in our analysis. These out-of-Qatar deaths may introduce differential ascertainment bias. However, the sensitivity analysis restricting the study to only Qataris showed similar results.

Since the magnitude of the Cox HR in presence of time-varying HR depends on the scale and distribution of losses to follow-up (censoring) even if the losses occur at random,^20^ the overall HRs in the post-acute analyses are skewed toward the earlier of times of follow-up rather than later times of follow-up. However, HRs were additionally provided for both early follow-up and late follow-up (Figure 2) to provide representative estimates for each of these durations.

The study was conducted in a specific national population consisting mainly of healthy working-age adults, and thus generalizability of the findings to other national populations is unknown. As an observational study, investigated cohorts were neither blinded nor randomized, so unmeasured or uncontrolled confounding cannot be excluded. Although matching covered key factors affecting risks of death or infection,^8,11-14^ it was not possible for other factors such as geography or occupation, for which data were unavailable. However, Qatar is essentially a city state and infection incidence was broadly distributed across neighborhoods. Nearly 90% of Qatar’s population are expatriates from over 150 countries, who come for employment.^8^ Nationality, age, and sex provide a powerful proxy for socio-economic status in this country.^8,11-14^ Nationality is strongly associated with occupation.^8,12-14^ The matching procedure used in this study was investigated in previous studies of different epidemiologic designs, and using control groups to test for null effects.^6,7,38-40^ These prior studies demonstrated at different times during the pandemic that this procedure balances differences in infection exposure to estimate vaccine effectiveness,^6,7,38-40^ suggesting that the matching strategy may also have overcome differences in mortality risk. Analyses were implemented on Qatar’s total population and large samples, perhaps minimizing the likelihood of bias.

In conclusion, forward displacement of deaths among persons with relatively short life expectancies is the main driver of COVID-19 mortality in Qatar. The mortality displacement particularly affected those more clinically vulnerable to severe COVID-19. Meanwhile, vaccination negated the pull forward effect by preventing early deaths. The findings do not diminish the importance of COVID-19 deaths in otherwise healthy persons or due to Long COVID, though there was no discernable effect for these deaths in Qatar’s population.

### Sources of support and acknowledgements

We acknowledge the many dedicated individuals at Hamad Medical Corporation, the Ministry of Public Health, the Primary Health Care Corporation, Qatar Biobank, Sidra Medicine, and Weill Cornell Medicine-Qatar for their diligent efforts and contributions to make this study possible.

The authors are grateful for institutional salary support from the Biomedical Research Program and the Biostatistics, Epidemiology, and Biomathematics Research Core, both at Weill Cornell Medicine-Qatar, as well as for institutional salary support provided by the Ministry of Public Health, Hamad Medical Corporation, and Sidra Medicine. The authors are also grateful for the Qatar Genome Programme and Qatar University Biomedical Research Center for institutional support for the reagents needed for the viral genome sequencing. The funders of the study had no role in study design, data collection, data analysis, data interpretation, or writing of the article.

Statements made herein are solely the responsibility of the authors.

## Data Availability

Data used in this study are the property of the Ministry of Public Health of Qatar and were provided to the researchers through a restricted-access agreement for preservation of confidentiality of patient data.

## Author contributions

JSF conceived the idea of this study. HC, JSF, and LJA conceived and designed the study analyses. HMK contributed to analysis plan. HC performed the statistical analyses and co-wrote the first draft of the article. LJA led the statistical analyses and co-wrote the first draft of the article. PT and MRH designed and conducted multiplex, RT-qPCR variant screening and viral genome sequencing. PVC designed mass PCR testing to allow routine capture of SGTF variants and conducted viral genome sequencing. HY, AAA, and HAK conducted viral genome sequencing. All authors contributed to data collection and acquisition, database development, discussion and interpretation of the results, and to the writing of the article. All authors have read and approved the final manuscript. Decision to publish the paper was by consensus among all authors.

## Supplementary Appendix

## Section S1. Further details on methods

### Population, data sources, and testing

Qatar has a population of about 3 million people.^1,2^ The population is unusually young and diverse in that only 9% of its residents are ≥50 years of age, and 89% are expatriates from over 150 countries.^1,2^ The population is dynamic because the vast majority of Qatar’s residents come here because of employment and leave the country after end of employment.^2^ Hosting of the World Cup 2022 and the years of infrastructure building that this has entailed contributed to a large surge and fluctuations in the population over the last few years. Nationality, age, and sex provide a powerful proxy for socio-economic status in this country,^2-6^ as nationality is strongly associated with occupation in this population.^2,4-6^ The national, federated, mortality database is managed by the Hamad Medical Corporation (HMC) since the start of the year 2020. HMC is the national public healthcare provider in the country. The database includes all death records, including both deaths occurring at healthcare facilities and elsewhere. The database includes also forensic deaths investigated by Qatar’s Ministry of Interior. The number of deaths recorded in Qatar since 2020 averaged at about 2,300 deaths per year.

Qatar launched its coronavirus disease 2019 (COVID-19) vaccination program in December of 2020 using the BNT162b2 and mRNA-1273 vaccines.^7^ Most of primary-series vaccination scale-up was implemented in 2021.^8,9^ Most of booster vaccination scale-up was implemented in 2022.^10,11^ Vaccination with the bivalent booster started towards the end of 2022, but it remains at low coverage. Nearly all individuals in the population were vaccinated free of charge in Qatar, rather than elsewhere.

Qatar’s national and universal public healthcare system uses the Cerner-system advanced digital health platform to track all electronic health record encounters of each individual in the country, including all citizens and residents registered in the national and universal public healthcare system. Registration in the public healthcare system is mandatory for citizens and residents.

The databases analyzed in this study are data-extract downloads from the Cerner-system that have been implemented on a regular (twice weekly) schedule since the onset of pandemic by the Business Intelligence Unit at HMC. At every download all tests, COVID-19 vaccinations, hospitalizations related to COVID-19, and all death records regardless of cause are provided to the authors through .csv files. These databases have been analyzed throughout the pandemic not only for study-related purposes, but also to provide policymakers with summary data and analytics to inform the national response.

Every health encounter in the Cerner-system is linked to a unique individual through the HMC Number that links all records for this individual at the national level. Databases were merged and analyzed using the HMC Number to link all records whether for testing, vaccinations, hospitalizations, and deaths. All deaths in Qatar are tracked by the public healthcare system. All COVID-19-related healthcare was provided only in the public healthcare system. No private entity was permitted to provide COVID-19-related healthcare. COVID-19 vaccination was also provided only through the public healthcare system. These health records were tracked throughout the COVID-19 pandemic using the Cerner system. This system has been implemented in 2013, before the onset of the pandemic. Therefore, we had all health records related to this study for the full national cohort of citizens and residents throughout the pandemic. This allowed us to follow each person over time.

Demographic details for every HMC Number (individual) such as sex, age, and nationality are collected upon issuing of the universal health card, based on the Qatar Identity Card, which is a mandatory requirement by the Ministry of Interior to every citizen and resident in the country.

Severe acute respiratory syndrome coronavirus 2 (SARS-CoV-2) testing in Qatar is done at a mass scale where close to 5% of the population are tested every week.^8,12^ All SARS-CoV-2 testing in any facility in this country is tracked nationally in one database, the national testing database. This database covers all testing in all locations and facilities throughout the country, whether public or private. Every polymerase chain reaction (PCR) test and an increasing proportion of the facility-based rapid antigen tests conducted in Qatar, regardless of location or setting, are classified on the basis of symptoms and the reason for testing (clinical symptoms, contact tracing, surveys or random testing campaigns, individual requests, routine healthcare testing, pre-travel, at port of entry, or other). Based on the distribution of the reason for testing up to October 31, 2022, most of the tests that have been conducted in Qatar were conducted for routine reasons, such as being travel-related. Greater than 70% of those diagnosed are also diagnosed not because of appearance of symptoms, but because of routine testing (Table 1).^8,12^ All testing results in the national testing database during follow-up in the present study were factored in the analyses of this study.

The first large omicron wave that peaked in January of 2022 was massive and strained the testing capacity in the country.^12,13^ Accordingly, rapid antigen testing was introduced to relieve the pressure on PCR testing. Implementation of this change in testing occurred quickly precluding incorporation of reason for testing in a proportion of the rapid antigen tests for several months.

While the reason for testing is available for all PCR tests, it is not available for all rapid antigen tests.

Rapid antigen test kits are available for purchase in pharmacies in Qatar, but outcome of home-based testing is not reported nor documented in the national databases. Since SARS-CoV-2-test outcomes are linked to specific public health measures, restrictions, and privileges, testing policy and guidelines stress medically supervised testing as the core testing mechanism in the population. While medically supervised testing is provided free of charge or at low subsidized costs, depending on the reason for testing, home-based rapid antigen testing is de-emphasized and not supported as part of national policy. There is no reason to believe that home-based testing could have differentially affected the followed matched cohorts to affect our results.

Further descriptions of the study population and these national databases were reported previously.^2,8,10,12,14^

### Comorbidity classification

Comorbidities were ascertained and classified based on the ICD-10 codes for chronic conditions as recorded in the electronic health record encounters of each individual in the Cerner-system national database that includes all citizens and residents registered in the national and universal public healthcare system. The public healthcare system provides healthcare to the entire resident population of Qatar free of charge or at heavily subsidized costs, including prescription drugs.

All encounters for each individual were analyzed to determine the comorbidity classification for that individual, as part of a recent national analysis to assess healthcare needs and resource allocation. The Cerner-system national database includes encounters starting from 2013, after this system was launched in Qatar. As long as each individual had at least one encounter with a specific comorbidity diagnosis since 2013, this person was classified with this comorbidity.

Individuals who have comorbidities but never sought care in the public healthcare system, or seek care exclusively in private healthcare facilities, were classified as individuals with no comorbidity due to absence of recorded encounters for them. This misclassification bias is not likely to affect the study results. The results for those more clinically vulnerable will not be affected, as this misclassification bias would have only resulted in a smaller cohort of these persons. This cohort was large enough for precise estimation of outcomes. As for those less clinically vulnerable, the misclassification bias could imply that some of them may have been more clinically vulnerable. However, this proportion is likely to be very small compared to the proportion of those with one or no comorbidity in the young population of Qatar. The effect on study outcomes is thus likely to be negligible.

### Classification of COVID-19 death

Classification of COVID-19 death followed World Health Organization (WHO) guidelines.^15^ Assessments were made by trained medical personnel independent of study investigators and using individual chart reviews, as part of a national protocol applied to every deceased patient since pandemic onset.

COVID-19 death was defined per WHO classification as “a death resulting from a clinically compatible illness, in a probable or confirmed COVID-19 case, unless there was a clear alternative cause of death that could not be related to COVID-19 disease (e.g. trauma), and there was no period of complete recovery between the COVID-19 illness and death. A death due to COVID-19 could not be attributed to another disease (e.g. cancer) and was counted independently of preexisting conditions suspected of triggering or increasing the risk of a severe course of COVID-19”. Detailed WHO criteria for classifying COVID-19 deaths can be found in the WHO technical report.^15^

## Section S2. Laboratory methods and variant ascertainment

### Real-time reverse-transcription polymerase chain reaction testing

Nasopharyngeal and/or oropharyngeal swabs were collected for polymerase chain reaction (PCR) testing and placed in Universal Transport Medium (UTM). Aliquots of UTM were: 1) extracted on KingFisher Flex (Thermo Fisher Scientific, USA), MGISP-960 (MGI, China), or ExiPrep 96 Lite (Bioneer, South Korea) followed by testing with real-time reverse-transcription PCR (RT-qPCR) using TaqPath COVID-19 Combo Kits (Thermo Fisher Scientific, USA) on an ABI 7500 FAST (Thermo Fisher Scientific, USA); 2) tested directly on the Cepheid GeneXpert system using the Xpert Xpress SARS-CoV-2 (Cepheid, USA); or 3) loaded directly into a Roche cobas 6800 system and assayed with the cobas SARS-CoV-2 Test (Roche, Switzerland). The first assay targets the viral S, N, and ORF1ab gene regions. The second targets the viral N and E-gene regions, and the third targets the ORF1ab and E-gene regions.

All PCR testing was conducted at the Hamad Medical Corporation Central Laboratory or Sidra Medicine Laboratory, following standardized protocols.

### Rapid antigen testing

Severe acute respiratory syndrome coronavirus 2 (SARS-CoV-2) antigen tests were performed on nasopharyngeal swabs using one of the following lateral flow antigen tests: Panbio COVID-19 Ag Rapid Test Device (Abbott, USA); SARS-CoV-2 Rapid Antigen Test (Roche, Switzerland); Standard Q COVID-19 Antigen Test (SD Biosensor, Korea); or CareStart COVID-19 Antigen Test (Access Bio, USA). All antigen tests were performed point-of-care according to each manufacturer’s instructions at public or private hospitals and clinics throughout Qatar with prior authorization and training by the Ministry of Public Health (MOPH). Antigen test results were electronically reported to the MOPH in real time using the Antigen Test Management System which is integrated with the national Coronavirus Disease 2019 (COVID-19) database.

**Table S1.**
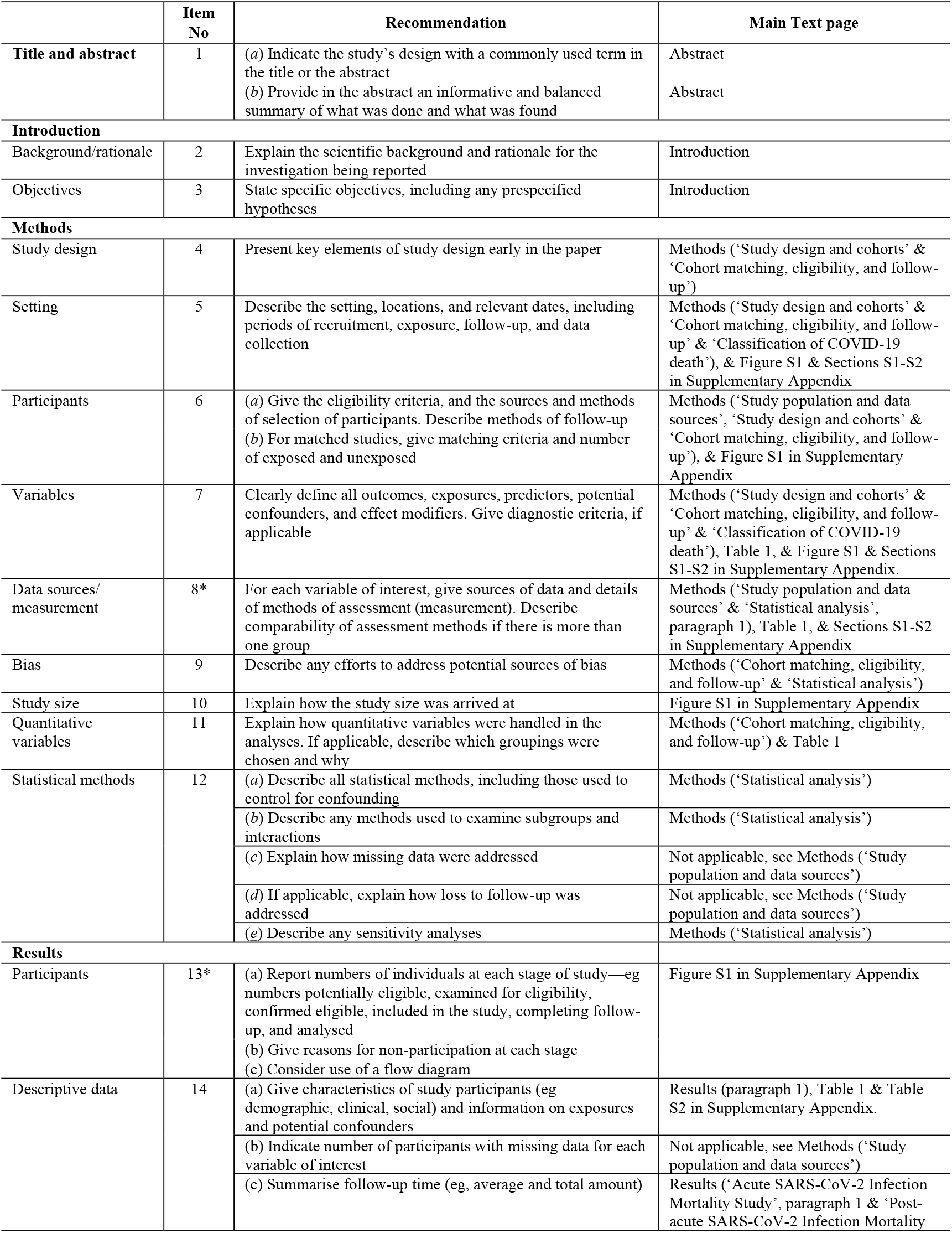
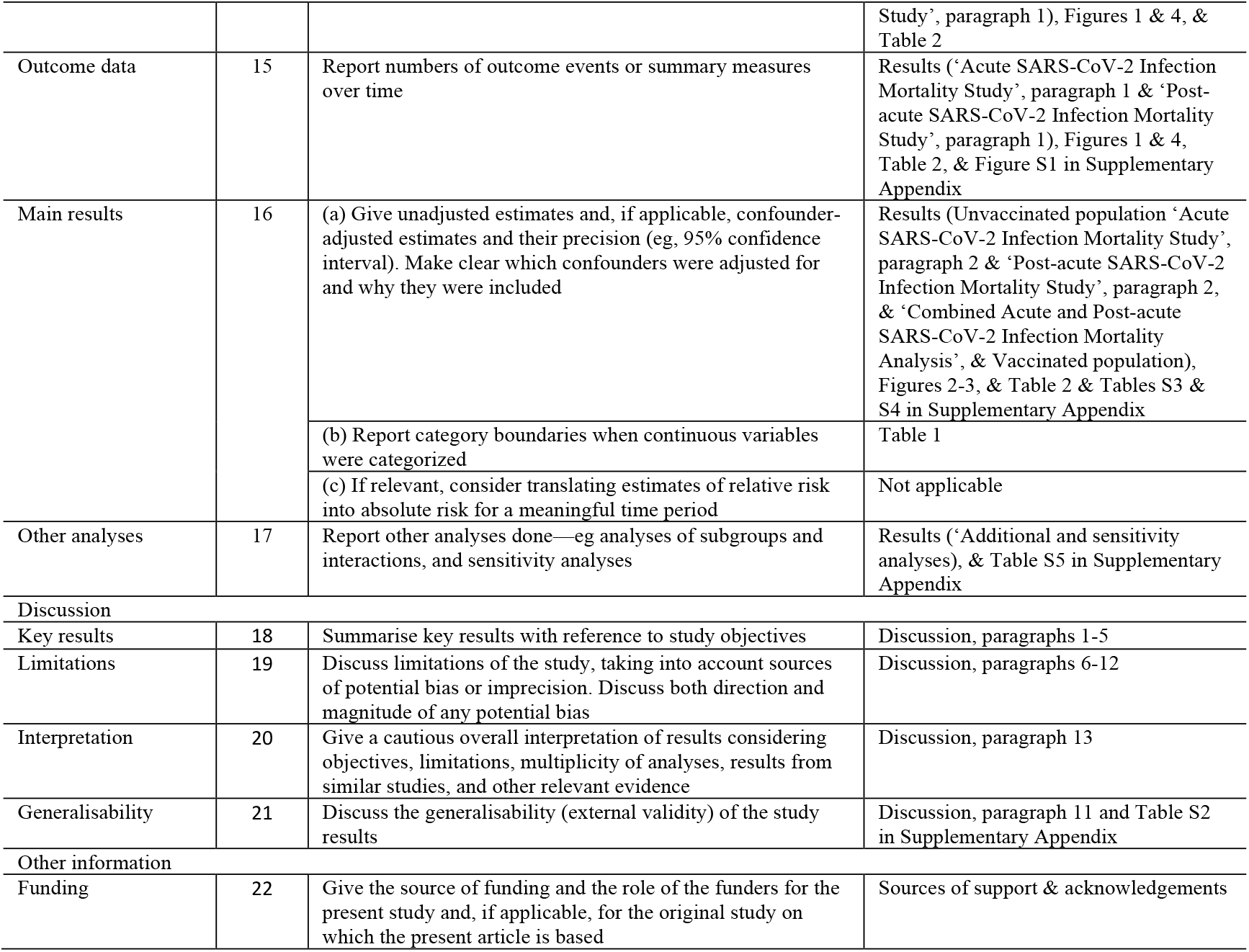
STROBE checklist for cohort studies.

**Figure S1.**
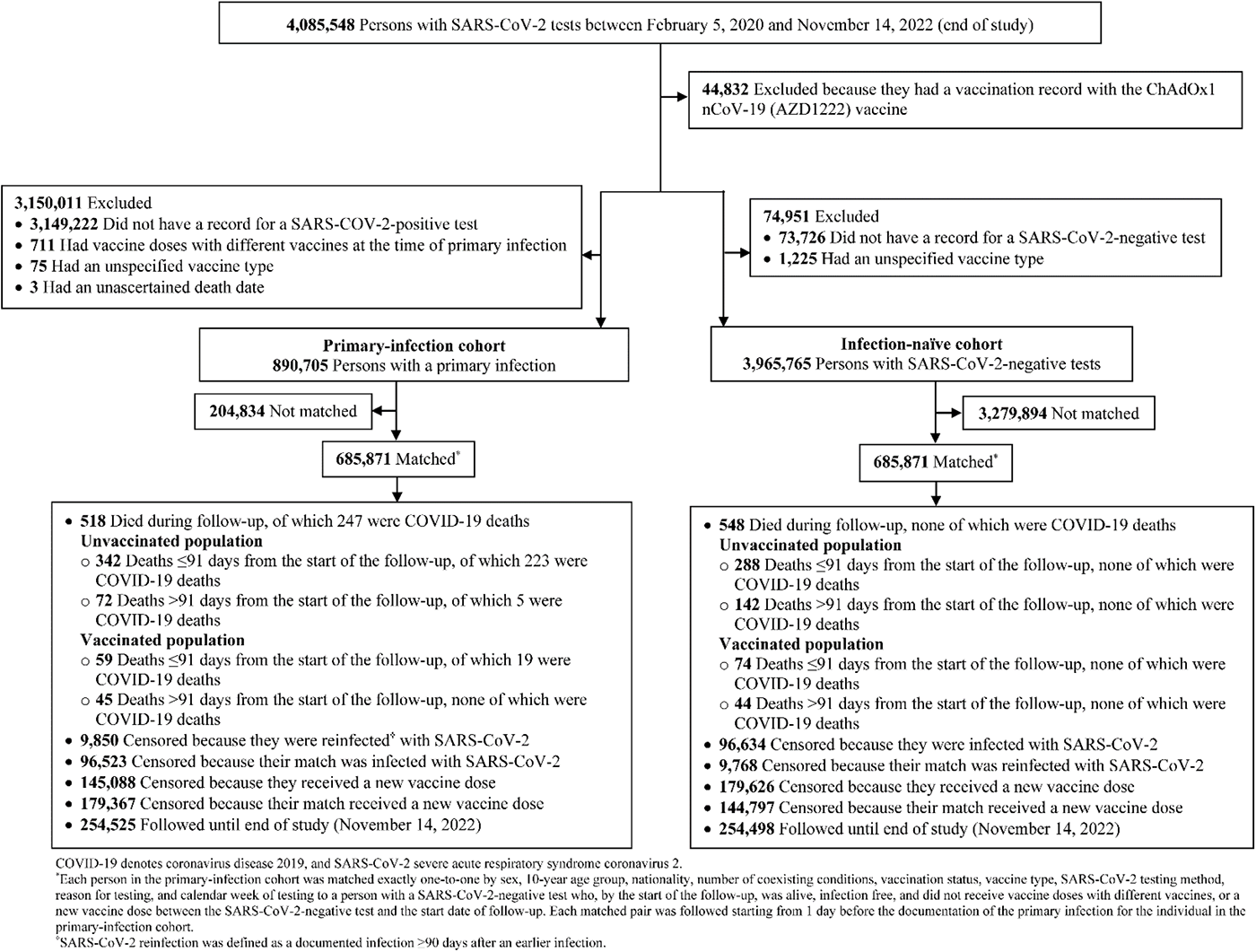
Cohort selection for investigating the risk of all-cause mortality in the primary-infection cohort relative to the infection-naive cohort.

**Table S2.** Representativeness of study participants.

| Category |  |
| --- | --- |
| Disease, problem, or condition under investigation | Incidence of all-cause death was investigated in the national cohort of SARS-CoV-2 infected persons compared to that in a reference national control cohort of infection-naïve persons by vaccination status, and in subgroups of persons who are less or more clinically vulnerable to severe COVID-19 given their age and number of coexisting conditions. |
| Special considerations related to |  |
| Sex and gender | A national, matched, retrospective, cohort study was conducted to compare incidence of all-cause mortality in the primary-infection cohort to that in the infection-naïve cohort in the acute phase of infection ($\leq 90$ days after primary infection) and in the post-acute phase ($> 90$ days after primary infection). Cohorts were matched exactly one-to-one by sex to control for differences in the risk of death by sex. |
| Age | Cohorts were matched exactly one-to-one by age to control for differences in the risk of death by age. |
| Race or ethnicity group | Cohorts were matched exactly one-to-one by nationality to control for potential differences in the risk of death by nationality. Nationality is associated with race and ethnicity in the population of Qatar. |
| Geography | Individual-level data on geography were not available, but Qatar is essentially a city state and infection incidence was broadly distributed across the country's neighborhoods/areas. Cohorts were matched exactly one-to-one by nationality to control for potential differences in the risk of death by nationality. Qatar has unusually diverse demographics in that 89% of the population are international expatriate residents coming from over 150 countries from all world regions. |
| Other considerations | Persons were matched exactly by number of coexisting conditions to control for differences in the risk of death by coexisting conditions. Persons were also matched exactly by vaccination status (number of doses) and mRNA vaccine type to control for the protective effect of vaccination. Persons were further matched exactly by testing method and by reason for testing to control for potential differences in testing modalities between cohorts. Persons were additionally matched by calendar week of testing. That is, matched pairs needed to have been tested for SARS-CoV-2 in the same calendar week to control for time-variable differences in mortality risk between cohorts and to ensure that matched pairs were present in Qatar in the same period. |
| Overall representativeness of this study | The study was based on the total population of Qatar and thus the study population is broadly representative of the diverse, by national background, but young total population of Qatar. While there could be differences in the risk of death by sex, age, nationality, number of coexisting conditions, vaccinations status, vaccine type, SARS-CoV-2 testing method, reason for testing, and calendar week of testing, cohorts were matched exactly by these factors to control for their potential impact on our estimates. |

SARS-CoV-2 denotes severe acute respiratory syndrome coronavirus 2.

**Table S3.** Adjusted hazard ratios for all-cause death by clinical vulnerability in the A) unvaccinated and B) vaccinated populations. 13.

| Epidemiological measures | Acute SARS-CoV-2 Infection Mortality Analysis |  | Post-acute SARS-CoV-2 Infection Mortality Analysis |  | Acute & Post-acute SARS-CoV-2 Infection Mortality Analysis |  |
| --- | --- | --- | --- | --- | --- | --- |
|  | Primary-infection cohort* | Control cohort* | Primary-infection cohort* | Control cohort* | Primary-infection cohort* | Control cohort* |
| A) Unvaccinated population |  |  |  |  |  |  |
| Less clinically vulnerable |  |  |  |  |  |  |
| Sample size | 415,596 | 415,596 | 285,006 | 284,984 | 415,596 | 415,596 |
| Total follow-up time (person-years) | 85,223 | 85,219 | 172,002 | 171,972 | 257,225 | 257,191 |
| Number of deaths during follow-up | 105 | 111 | 33 | 41 | 138 | 152 |
| Incidence rate of death (per 1,000 person-years; 95% CI) | 1.23 (1.02 to 1.49) | 1.30 (1.08 to 1.57) | 0.19 (0.14 to 0.27) | 0.24 (0.18 to 0.32) | 0.54 (0.45 to 0.63) | 0.59 (0.50 to 0.69) |
| Unadjusted hazard ratio for death (95% CI) | 0.95 (0.73 to 1.25) |  | 0.78 (0.49 to 1.24) |  | 0.91 (0.72 to 1.14) |  |
| Adjusted hazard ratio for death (95% CI)<sup>†</sup> | 0.94 (0.72 to 1.24) |  | 0.77 (0.48 to 1.24) |  | 0.90 (0.71 to 1.14) |  |
| More clinically vulnerable |  |  |  |  |  |  |
| Sample size | 45,024 | 45,024 | 30,483 | 30,522 | 45,024 | 45,024 |
| Total follow-up time (person-years) | 9,044 | 9,052 | 15,357 | 15,347 | 24,401 | 24,399 |
| Number of deaths during follow-up | 237 | 177 | 39 | 101 | 276 | 278 |
| Incidence rate of death (per 1,000 person-years; 95% CI) | 26.21 (23.07 to 29.76) | 19.55 (16.88 to 22.66) | 2.54 (1.86 to 3.48) | 6.58 (5.41 to 8.00) | 11.31 (10.05 to 12.73) | 11.39 (10.13 to 12.82) |
| Unadjusted hazard ratio for death (95% CI) | 1.31 (1.08 to 1.59) |  | 0.39 (0.27 to 0.57) |  | 0.99 (0.84 to 1.17) |  |
| Adjusted hazard ratio for death (95% CI)<sup>†</sup> | 1.34 (1.11 to 1.63) |  | 0.37 (0.25 to 0.54) |  | 1.00 (0.85 to 1.19) |  |
| B) Vaccinated population |  |  |  |  |  |  |
| Less clinically vulnerable |  |  |  |  |  |  |
| Sample size | 186,894 | 186,894 | 109,504 | 109,500 | 186,894 | 186,894 |
| Total follow-up time (person-years) | 33,603 | 33,602 | 47,288 | 47,286 | 80,891 | 80,888 |
| Number of deaths during follow-up | 16 | 18 | 12 | 7 | 28 | 25 |
| Incidence rate of death (per 1,000 person-years; 95% CI) | 0.48 (0.29 to 0.78) | 0.54 (0.34 to 0.85) | 0.25 (0.14 to 0.45) | 0.15 (0.07 to 0.31) | 0.35 (0.24 to 0.50) | 0.31 (0.21 to 0.46) |
| Unadjusted hazard ratio for death (95% CI) | 0.89 (0.45 to 1.74) |  | 1.71 (0.67 to 4.35) |  | 1.12 (0.65 to 1.92) |  |
| Adjusted hazard ratio for death (95% CI)<sup>†</sup> | 1.08 (0.51 to 2.29) |  | 2.00 (0.68 to 5.84) |  | 1.33 (0.72 to 2.46) |  |
| More clinically vulnerable |  |  |  |  |  |  |
| Sample size | 38,357 | 38,357 | 22,456 | 22,442 | 38,357 | 38,357 |
| Total follow-up time (person-years) | 6,823 | 6,821 | 9,233 | 9,223 | 16,056 | 16,044 |
| Number of deaths during follow-up | 43 | 56 | 33 | 37 | 76 | 93 |
| Incidence rate of death (per 1,000 person-years; 95% CI) | 6.30 (4.67 to 8.50) | 8.21 (6.32 to 10.67) | 3.57 (2.54 to 5.03) | 4.01 (2.91 to 5.54) | 4.73 (3.78 to 4.93) | 5.80 (4.73 to 7.10) |
| Unadjusted hazard ratio for death (95% CI) | 0.77 (0.52 to 1.14) |  | 0.89 (0.56 to 1.42) |  | 0.82 (0.60 to 1.11) |  |
| Adjusted hazard ratio for death (95% CI)<sup>†</sup> | 0.64 (0.41 to 1.02) |  | 0.96 (0.56 to 1.66) |  | 0.76 (0.53 to 1.07) |  |

CI denotes confidence interval.
\*Each person in the primary-infection cohort was matched exactly one-to-one by sex, 10-year age group, nationality, number of coexisting conditions, vaccination status, vaccine type, SARS-CoV-2 testing method, reason for testing, and calendar week of testing to a person with a SARS-CoV-2-negative test who was alive, infection free, and did not receive a new vaccine dose at the start of follow-up.
<sup>†</sup>Cox regression analysis adjusted for sex, 10-year age group, 10 nationality groups, number of coexisting conditions, vaccination status, vaccine type, SARS-CoV-2 testing method, reason for testing, and calendar week of testing.

**Table S4.** Adjusted hazard ratio for all-cause death in the full matched primary-infection and infection-naïve cohorts, including both unvaccinated and vaccinated persons, over the entire time of follow-up, that is combining the acute and post-acute phases. 14.

| Epidemiological measures | Primary-infection cohort* | Control cohort* |
| --- | --- | --- |
| **SARS-CoV-2 Infection Mortality** |  |  |
| Sample size | 685,871 | 685,871 |
| Total follow-up time (person-years) | 378,573 | 378,523 |
| Number of deaths during follow-up | 518 | 548 |
| Incidence rate of death (per 1,000 person-years; 95% CI) | 1.37 (1.26 to 1.49) | 1.45 (1.33 to 1.57) |
| Unadjusted hazard ratio for death (95% CI) | 0.94 (0.84 to 1.06) |  |
| Adjusted hazard ratio for death (95% CI)<sup>†</sup> | 0.95 (0.84 to 1.07) |  |

CI denotes confidence interval.
\*Each person in the primary-infection cohort was matched exactly one-to-one by sex, 10-year age group, nationality, number of coexisting conditions, vaccination status, vaccine type, SARS-CoV-2 testing method, reason for testing, and calendar week of testing to a person with a SARS-CoV-2-negative test who, by the start of the follow-up, was alive, infection free, and did not receive vaccine doses with different vaccines, or a new vaccine dose between the SARS-CoV-2-negative test and the start date of follow-up.
<sup>†</sup>Cox regression analysis adjusted for sex, 10-year age group, 10 nationality groups, number of coexisting conditions, vaccination status, vaccine type, SARS-CoV-2 testing method, reason for testing, and calendar week of testing.

**Table S5.** Adjusted hazard ratio for all-cause death in the matched^*^ primary-infection and infection-naïve cohorts for each of unvaccinated and vaccinated persons in A) the model including an interaction term between study cohorts and vaccination status and B) the main analysis.

| Adjusted hazard ratio for death in the primary infection cohort relative to the infection-naïve cohort (95% CI) | Unvaccinated | Vaccinated |
| --- | --- | --- |
| **A) Model including an interaction term between study cohorts and vaccination status<sup>†</sup>** |  |  |
| Acute SARS-CoV-2 Infection Mortality Analysis | 1.19 (1.01 to 1.39) | 0.74 (0.50 to 1.10) |
| Post-acute SARS-CoV-2 Infection Mortality Analysis | 0.50 (0.37 to 0.67) | 1.11 (0.69 to 1.78) |
| **B) Main analysis restricted to Qataris only<sup>‡</sup>** |  |  |
| Acute SARS-CoV-2 Infection Mortality Analysis | 1.14 (0.83 to 1.56) | 0.74 (0.44 to 1.27) |
| Post-acute SARS-CoV-2 Infection Mortality Analysis | 0.46 (0.29 to 0.74) | 1.52 (0.82 to 2.82) |

CI denotes confidence interval.
\*Each person in the primary-infection cohort was matched exactly one-to-one by sex, 10-year age group, nationality, number of coexisting conditions, vaccination status, vaccine type, SARS-CoV-2 testing method, reason for testing, and calendar week of testing to a person with a SARS-CoV-2-negative test who, by the start of the follow-up, was alive, infection free, and did not receive vaccine doses with different vaccines, or a new vaccine dose between the SARS-CoV-2-negative test and the start date of follow-up.
<sup>†</sup>Cox regression analysis adjusted for sex, 10-year age group, 10 nationality groups, number of coexisting conditions, vaccine type, SARS-CoV-2 testing method, reason for testing, and calendar week of testing.
<sup>‡</sup>Cox regression analysis adjusted for sex, 10-year age group, number of coexisting conditions, vaccination status, vaccine type, SARS-CoV-2 testing method, reason for testing, and calendar week of testing.

